# Development and evaluation of a machine learning-based in-hospital COvid-19 Disease Outcome Predictor (CODOP): a multicontinental retrospective study

**DOI:** 10.1101/2021.09.20.21263794

**Authors:** Riku Klén, Disha Purohit, Ricardo Gómez-Huelgas, José Manuel Casas-Rojo, Juan Miguel Antón Santos, Jesús Millán Núñez-Cortés, Carlos Lumbreras, José Manuel Ramos-Rincón, Pablo Young, Juan Ignacio Ramírez, Estela Edith Titto Omonte, Rosmery Gross Artega, Magdy Teresa Canales Beltrán, Pascual Valdez, Florencia Pugliese, Rosa Castagna, Nico Funke, Benjamin Leiding, David Gómez-Varela

**Affiliations:** Turku PET Centre, University of Turku, Finland; Max Planck Institute of Experimental Medicine, Göttingen, Germany; Systems Biology of Pain, Division of Pharmacology & Toxicology, Department of Pharmaceutical Sciences, University of Vienna, Austria; Internal Medicine Department, Regional University Hospital of Málaga, Biomedical Research Institute of Málaga (IBIMA), University of Málaga (UMA), Málaga, Spain; Internal Medicine Department, Infanta Cristina University Hospital, Parla, Madrid, Spain; Internal Medicine Department, Gregorio Marañón University Hospital, Madrid, Spain; Internal Medicine Department, 12 de Octubre University Hospital, Madrid, Spain; Internal Medicine Department, General University Hospital of Alicante, Alicante Institute for 22 Health and Biomedical Research (ISABIAL), Miguel Hernández Elche Unit, Alicante, Spain; Clinical Medicine service, Hospital Británico of Buenos Aires, Buenos Aires, Argentina; Internal Medicine Service, Hospital Santa Cruz - Caja Petrolera de Salud, Santa Cruz de la Sierra, Bolivia; Hospital of San Juan de Dios, Epidemiology Unit, Santa Cruz, Bolivia; Instituto Hondureño of social security, Hospital Honduras Medical Centre, Tegucigalpa, Honduras; Hospital Vélez Sarsfield, Buenos Aires, Argentina; Institute for Software and Systems Engineering, TU Clausthal, Clausthal, Germany

## Abstract

**Background:** More contagious SARS-CoV-2 virus variants, breakthrough infections, waning immunity, and sub-optimal rates of COVID-19 vaccination account for a new surge of infections leading to record numbers of hospitalizations and deaths in several European countries. This is a particularly concerning scenario for resource-limited countries, which have a lower vaccination rate and fewer clinical tools to fight against the next pandemic waves. There is an urgent need for clinically valuable, generalizable, and parsimonious triage tools assisting the appropriate allocation of hospital resources. We aimed to develop and extensively validate CODOP, a machine learning-based tool for accurately predicting the clinical outcome of hospitalized COVID-19 patients.

**Methods:** CODOP was built using modified stable iterative variable selection and linear regression with lasso regularisation. To avoid generalization problems, CODOP was trained and tested with three time-sliced and geographically distinct cohorts encompassing 40 511 blood-based analyses of COVID-19 patients from more than 110 hospitals in Spain and the USA during 2020-21. We assessed the discriminative ability of the model using the Area Under the Receiving Operative Curve (AUROC) as well as horizon and Kaplan-Meier risk stratification analyses. To reckon the fluctuating pressure levels in hospitals through the pandemic, we offer two online CODOP calculators suited for undertriage or overtriage scenarios. We challenged their generalizability and clinical utility throughout an evaluation on a cohort of patients hospitalized in five hospitals from three Latin American countries.

**Findings:** CODOP uses 12 clinical parameters commonly measured at hospital admission and associated with the pathophysiology of COVID-19. CODOP reaches high discriminative ability up to nine days before clinical resolution (AUROC: 0·90-0·96, 95% CI 0·879-0·970), it is well calibrated, and it enables an effective dynamic risk stratification during hospitalization. The two CODOP online calculators demonstrate their potential for triage decisions when challenged with the distinctive Latin American evaluation cohorts (73-100% sensitivity and 84-100% specificity).

**Interpretation:** The high predictive performance of CODOP in geographically disperse patient cohorts and the easiness-of-use, strongly suggest its clinical utility as a global triage tool, particularly in resource-limited countries.

**Funding:** The Max Planck Society.

**Research in context:** *Evidence before this study:* We have searched PubMed for articles about the existence of in-hospital COVID-19 mortality predictive models, using the search terms “coronavirus”, “COVID-19”, “risk”, “death”, “mortality”, and “prediction”, focusing on studies published between March 1, 2020 and 31 August, 2021. The studies we identified generally used small-medium size cohorts of patients that are geographically restricted to small regions of the developed world (many times, to the same city). We haven’t found studies that challenged their models in extended cohorts of patients from very distinct health system populations, particularly from resource-limited countries. Further, most of the previous models are rigid by not acknowledging the fluctuating availability of hospital resources during the pandemic (e.g., beds, oxygen supply). These and other limitations have been pointed out by expert reviews indicating that published in-hospital COVID-19 mortality predictive models are subject to high risk of bias, report an over-optimistic performance, and have limited clinical value in assisting daily triage decisions. A parsimonious, accurate and extensively validated model is yet to be developed.

*Added value of this study:* We analysed clinical data from different cohorts totalling 21 607 COVID-19 patients treated in more than 110 hospitals in Spain and the USA during three different pandemic waves extending from February 2020 to April 2021. The new CODOP in-hospital mortality prediction model is based on 11 blood biochemistry parameters (representing main biological pathways involved in the pathogenesis of SARS-CoV-2) plus Age, all of them commonly measured upon hospitalization. CODOP accurately predicted mortality risk up to nine days before clinical resolution (AUROC: 0·90-0·96, 95% CI 0·879-0·970), it is well calibrated, and it enables an effective dynamic risk stratification during hospitalization. We offer two online CODOP calculator subtypes (https://gomezvarelalab.em.mpg.de/codop/) tailored to overtriage and undertriage scenarios. The online calculators were able to reach the desired prediction performance in five independent evaluation cohorts gathered in hospitals of three Latin American countries from March 7th 2020 to June 7th 2021.

*Implications of all the available evidence:* We present here a highly accurate, parsimonious and extensively validated COVID-19 in-hospital mortality prediction model, derived from working with the largest number and the most geographically extended representation of patients and health systems to date. The rigorous analytical methods, the generalizability of the model in distinct world regions, and its flexibility to reckon with the changing availability of hospital resources point to CODOP as a clinically useful tool potentially improving the outcome prediction and the management of COVID-19 hospitalized patients.

## Introduction

Since the first reported case in Wuhan at the end of 2019, COVID-19 has exerted extreme pressure on hospitals throughout the globe. The World Health Organization (WHO) estimated the pandemic as the direct cause of more than 5 million deaths. Despite positive data showing a decrease in hospitalizations and deaths among vaccinated people, warning signs forecast a scenario with health systems under severe strains leading to a bigger number of COVID-19 related deaths. The appearance of viral variants that are more contagious and that carry a higher risk of hospitalization,^1^ the waning of the immune protection, the significant amount of infections in vaccinated individuals (breakthrough infections) together with their ability to transmit the virus, and the slow and unequal rollout of vaccines worldwide, support recent models showing that a vaccine-alone exit strategy will likely not be sufficient to contain further outbreaks and their consequences.^2^ At the time of submission of this study, many European countries are reaching record-high numbers of infections, hospitalizations and deaths. This new pandemic wave depicts a worrisome prospect for resource-limited countries with similar or lower vaccination rates and with fewer clinical tools.

Prediction models that estimate the risk of death in hospitalized COVID-19 patients could be valuable both to clinicians and patients by assisting medical staff to stratify treatment strategy and by planning for appropriate allocation of limited resources. Thus, numerous models have been developed to assist triage decisions of hospitalized COVID-19 patients. However, independent evaluations have pointed out their lack of generalizability and their limited clinical use^3, 4^ due to causes belonging to the “dataset shift” problem.^5^ Moreover, the heterogeneity of the host-pathogen interaction (what results in more than 60 disease subtypes of COVID-19^6^) together with the fast evolution of the pandemic makes COVID-19 outcome prediction a challenging endeavour, especially if a profound evaluation using patient cohorts from geographically distinct regions is not performed.

To address this need, we used the largest and the most geographically extended patient’s dataset to date for developing and extensively validating a simple but yet clinically useful machine learning-based online model for doctors to predict mortality in COVID-19 patients at any time during hospitalization. To assist the real clinical needs during different pandemic scenarios we offer two predictor subtypes suited for undertriage and overtriage situations (https://gomezvarelalab.em.mpg.de/codop/).

The collective effort presented here unveils the power of machine learning for helping clinicians and patients in this pandemic. Based on its easiness to use and its generalizability among geographically very distinct patient cohorts, we aim for CODOP to become a useful triage tool, particularly in resource-limited countries.

## Methods

### Patient cohorts

The training and two test cohorts (test 1 and test 2) of this study are based on the SEMI (Sociedad Espanola de Medicina Interna) COVID-19 Registry.^7^ It is an ongoing multicentre nationwide cohort of consecutive patients hospitalized for COVID-19 across different Spanish regions (109 hospitals). Eligibility criteria were age ≥ 18 years, confirmed diagnosis of COVID-19, defined as a positive result on real-time reverse-transcription-polymerase-chain-reaction (RT-PCR) for the presence of SARS-CoV-2 in nasopharyngeal swab specimens or sputum samples, first hospital admission for COVID-19, and hospital discharge or in-hospital death.^7^ The SEMI-COVID-Registry was approved by the Provincial Research Ethics Committee of Málaga (Spain) and the Institutional Research Ethics Committees of each participating hospital.

The test cohort from New York is based on the study from Del Valle et. al.^8^ consisting of 2 021 COVID-19 patients hospitalized in the Mount Sinai Health System in New York City between March 21st and April 28th, 2020.

The evaluation cohorts used in the evaluation of the two online CODOP subtypes were provided by Honduras Medical Centre (45 patients, Tegucigalpa, Honduras), Hospital Santa Cruz Caja Petrolera de Salud (30 patients, Santa Cruz de la Sierra, Bolivia), Hospital San Juan de Dios (93 patients, Santa Cruz, Bolivia), Hospital Vélez Sarsfield (100 patients, Buenos Aires, Argentina), and Hospital Británico de Buenos Aires (150 patients, Buenos Aires, Argentina). The released of anonymized clinical data of all patients with COVID-19 used in this study has been reviewed by the institutional ethical review boards for each institution participating in this study.

### Predictors and outcomes

We included patient characteristics and blood test values (see Supplementary Table 1) that were present in all training and test cohorts, measured at different times during hospitalization, as potential predictors. We limited our potential predictors to variables that had less than 40% missing values. The percentage of missing values is listed in Supplementary Table 1. Most of the variables have less than 5% of missing values. Missing values were imputed in all datasets using the mean value of original variables in the training cohort. We trained a binary classification model in which the outcome is patient mortality: 1, if the patient was deceased, or 0, if discharged.

For each cohort, the subjects were divided into two groups based on their survival status. The normality of each numerical variable in the groups was tested with the Shapiro-Wilk normality test. None of the variables was normally distributed. For each variable statistical difference was tested between the two groups with the Wilcoxon rank-sum test for numerical variables and with the chi-squared test for categorical variables. The obtained P-values were adjusted for multiple testing by Benjamini-Hochberg Procedure.

### CODOP development

CODOP was built using modified stable iterative variable selection (SIVS)^9^ and linear regression with least absolute shrinkage and selection operator (lasso) regularisation.^10^ In model building only the training cohort was used and models were built using 10-fold cross-validation. In the feature selection stage of SIVS, 100 models were built and for each model selected variables were recorded. For reducing the number of features to as few as possible (therefore, increasing the easiness of use of CODOP), we tuned the weighting function in SIVS (called variable importance scoring) so that only features occurring in all of the 100 models were selected for the final model building stage. This method has shown to be very efficient, especially when the ratio of positive and negative outcomes is imbalanced^11^. Lasso models were built in R^12^ (version 3.6.0) package glmnet^10^ (version 4.1-1). All predictions were done blinded to the final clinical outcome. For converting numeric prediction into binary prediction, Youden’s J statistic was used.^13^ For building the two online CODOP subtypes we used alternative thresholds, which were selected to be the largest threshold value in the training cohort with a sensitivity of 95% for CODOP-Ovt and specificity of 95% for CODOP-Unt. Calibration plots were created with R package caret^14^ (version 6.0-86). Survival analysis was performed using univariable Cox proportional hazards regression model.^15^ Survival analysis and Kaplan-Meyer plots were produced with R packages survival^16^ (R package version 3.2-11) and survminer^17^ (R package version 0.4.9). For horizon analyses, the data were considered separately for survival time of one to nine days.

### Benchmarking

To evaluate the performance of CODOP we used three benchmark methods: COPE^18^, model by Zhang et al.^19^, and a univariable model. COPE model is a linear regression model, which uses variables age, respiratory rate, C-reactive protein, lactic dehydrogenase, albumin, and urea. Zhang et al. model is a logistic regression model, which uses variables age, sex, neutrophil count, lymphocyte, platelet, C-reactive protein, and creatinine. From the different models described in Zhang et al., model DL for prediction of death (Supplementary Table 2 of Zhang et al.) was used for benchmarking purposes. Univariable analysis was performed in the training dataset for all variables. The best univariable model was selected based on the average ranking of AUROC, accuracy, sensitivity and specificity. Different models were evaluated using four evaluation metrics: area under receiver operating curves (AUROC), accuracy, sensitivity, and specificity. The metrics were calculated using R packages pROC^20^ (version 1.17.0.1) and caret^14^ (R package version 6.0-86).

### Online evaluation

Five different Latin American hospitals provided the values for the 12 features used by CODOP that were measured in patients at two different time points between March 7th 2020 and June 7th 2021: during the time of hospitalization, and the worst values measured during hospitalization. The former datasets were used for calculating AUROC, calibration curves, and confusion matrices. Both times points were used for performing horizon analysis and risk-stratification. All predictions were done blinded to the final clinical outcome.

## Results

### CODOP development, performance and benchmark

We developed CODOP following a multistep process (Figure 1) using a training dataset with measurements of 20 features (18 blood biochemical parameters plus Age and Sex; Supplementary Table 1) routinely measured during admission on 15 902 COVID-19 patients hospitalized in 109 Spanish healthcare centres during the first COVID-19 wave that occurred in Spain between February 5th and July 6th 2020 (SEMI-COVID-19 Network database^7^).

**Figure 1.**
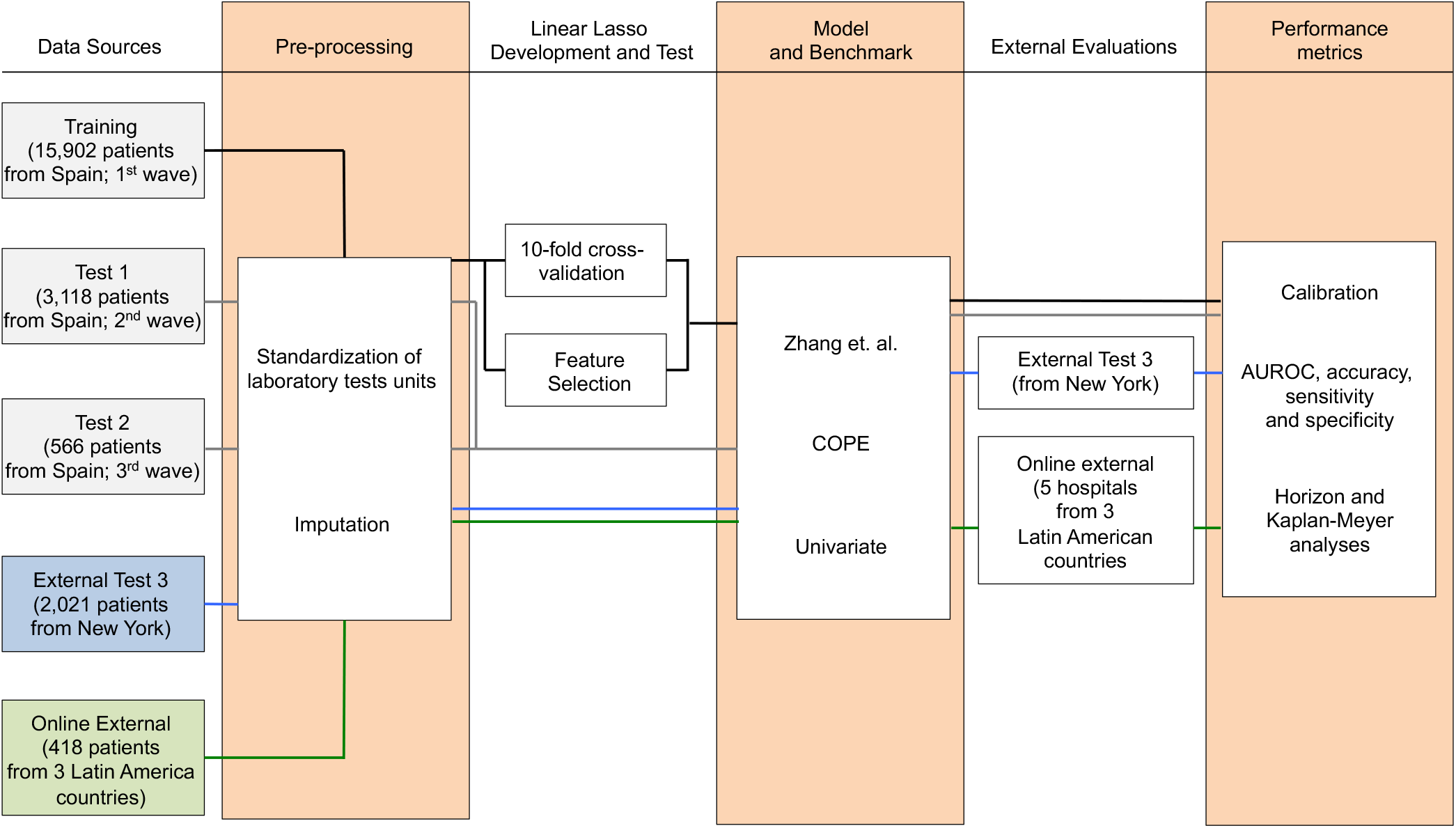
Flowchart depicting the different patient cohorts used in this study and the steps followed during the development, test, and independent evaluation of CODOP.

As a first step, data pre-processing included standardization of the laboratory tests units and imputation of the missing test values, which is characteristic for real-world clinical practice (Supplementary Table 1). Using linear Lasso, 10-fold cross-validation and SIVS we obtained a final CODOP model using 11 blood biochemical parameters plus Age (Supplementary Table 2 and Supplementary Figure 1). Detail analysis indicated that elevated values of Age, neutrophils, C-reactive protein, creatinine, lactate dehydrogenase, serum sodium, serum potassium, glucose and D-dimer, and reduced values of platelets, eosinophils and monocytes were positively correlated with in-hospital death, respectively (Supplementary Table 3).

Next, we benchmarked the performance of CODOP, using the same training dataset, against the predictor developed by Zhang et. al.^19^, against the predictor COPE^18^, and against Age (as the univariable feature with more predictive power; Supplementary Table 4). The two prognostic models were selected based on the availability of the model’s details and their use of blood-based features. CODOP showed a superior discriminative ability in predicting in-hospital mortality (area under the receiver operating curves or AUROC: 0·889, 95% CI 0·885-0·894; Figure 2A) reaching 0·84% and 0·78% sensitivity and specificity, respectively (Supplementary Table 5). In addition, CODOP has better calibration for all the different risk groups as reflected by a lower RMSE value (Figure 2B and Supplementary Table 6). A detailed inspection of the calibration curves shows that the predictor published by Zhang et al. underestimated the probability of death for low-risk patients and overestimates the probability of death for high-risk patients. On the other side, while COPE underestimates the probability of death for all risk groups, Age showed a clear overestimation (Figure 2B).

**Figure 2.**
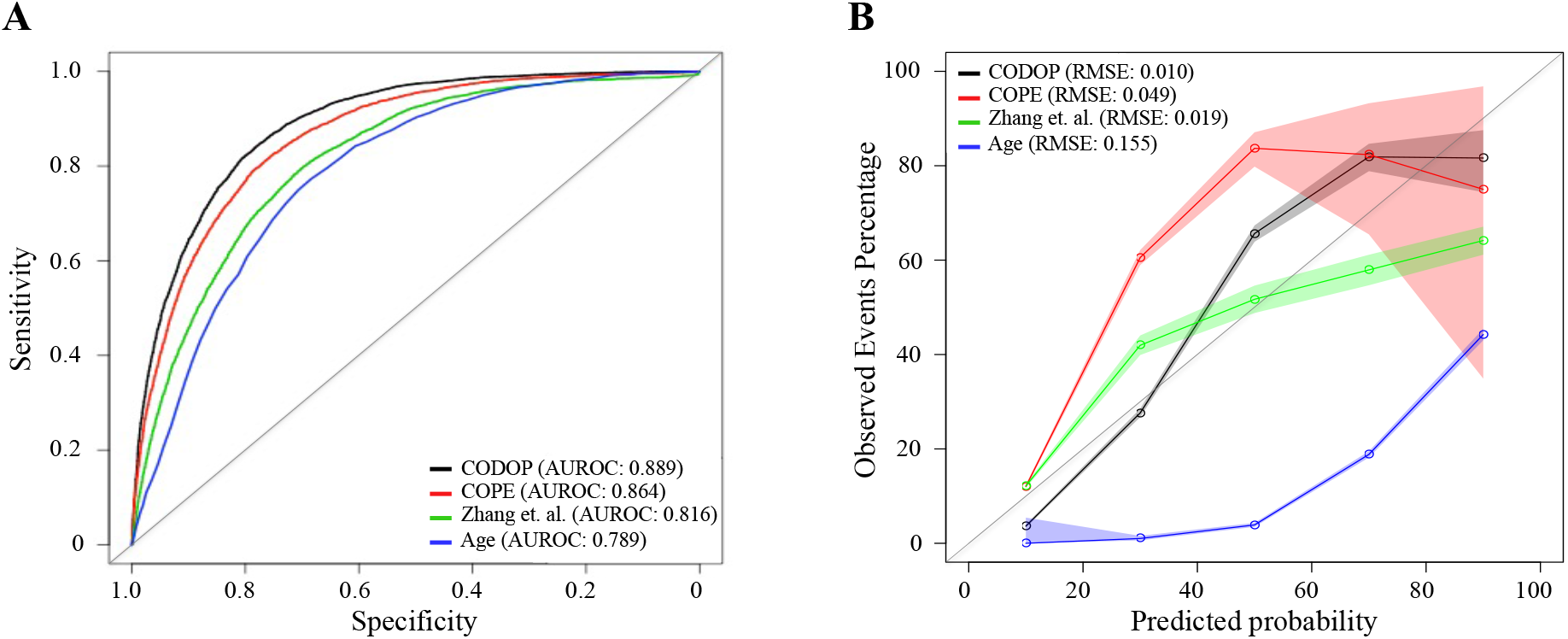
Discriminatory ability (using area under the receiver operating curves or AUROC; A) and calibration curves (B) for CODOP, COPE, Zhang et al., and Age in the training dataset.

### CODOP testing with independent and external cohorts

The size, demographic diversity (in terms of age, gender, ethnicity and comorbidities; see Table 1 of Rojo et. al. ^7^), and geographical spread of the training dataset, suggest the generalizability of the predictions made by CODOP. To challenge this, we investigated the discriminative ability and calibration of CODOP in three independent test cohorts.

On the one side, we used two time-sliced cohorts with COVID-19 patients hospitalized during the second and third COVID-19 waves that occurred in Spain between July 7th and December 6th 2020 (Test 1; 3 118 patients) and between December 7th 2020 and March 31st 2021 (Test 2; 566 patients). Notably, ROC and calibration curves show that the performance metrics are preserved in these two cohorts (Supplementary Figure 2, Supplementary Table 5 and Supplementary Table 6). Furthermore, the generalizability of CODOP was also demonstrated on a separate test cohort (External Test 3) consisting of 2 021 COVID-19 patients hospitalized in the Mount Sinai Health System in New York City between March 21st and April 28th, 2020.^8^ These results strongly suggest that CODOP is not influenced by the inherent changes occurring during the pandemic (e.g. the appearance of different virus variants, the used of more tailored clinical interventions, etc.). Of note, the percentage of deaths among these test cohorts is similar (Supplementary Table 7), ruling out an artefactual influence due to the time-sliced nature of these cohorts. Finally, CODOP overperformed both of the benchmarked predictors and Age in the three test cohorts (Supplementary Figure 2, Supplementary Table 5, and Supplementary Table 6), suggesting that it captures key biomarkers involved in the physiological deterioration of COVID-19 hospitalized patients.

### Estimation of fixed prediction horizons and dynamic risk-stratification

Many patients of the different cohorts had multiple blood samples taken during their hospitalization. We compare the discriminative ability of CODOP at a fixed time before the clinical resolution using the training cohort. On average, CODOP predicted the outcome of all patients nine days in advance with an average sensitivity (at a fixed specificity of 75%) and AUROC values higher than 90% (Figure 3A and Supplementary Table 8, respectively). In comparison, CODOP maintained a stable sensitivity along the nine days horizon time significantly outperforming (P<0.01, paired two-sided T-test) the other benchmarked predictors.

**Figure 3.**
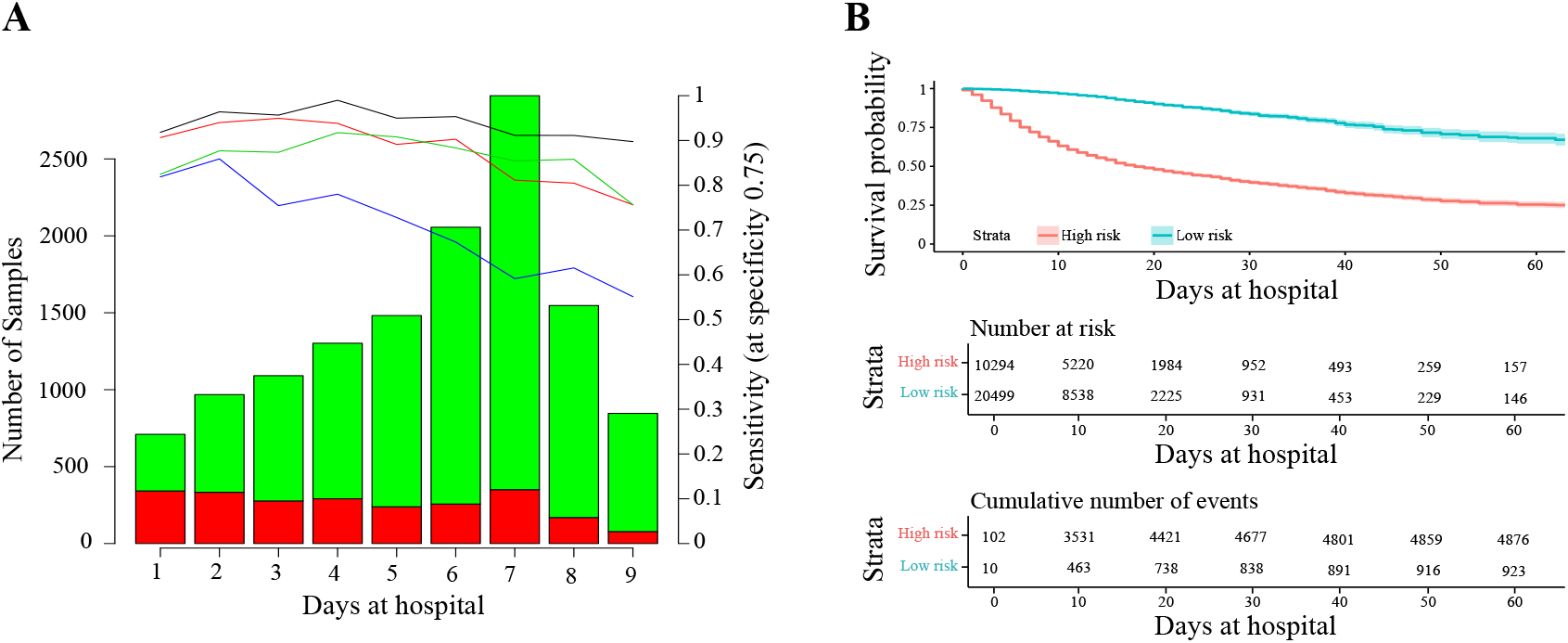
Horizon analysis (A) and survival analysis (B) in the training dataset. In the horizon plot x-axis represents the number of days at the hospital before clinical resolution, the bar plot is for the number of samples (the green colour is for survival and red for death), and lines are for sensitivity when the specificity was fixed at 75% in the training cohort (the black line is CODOP, the red line is COPE, the green line is Zhang et al., and the blue line is Age).

Next, we demonstrated that CODOP enables a continuous stratification of patients into a high-risk group over the course of the hospitalization, as patients with a higher CODOP score assigned were more likely to die over time (Figure 3B). We obtained similar stratification results when using the three test cohorts (Supplementary Figure 3). Hence, CODOP represents an early and dynamic warning tool in the clinical status of COVID-19 patients.

### Multinational evaluation of an online CODOP predictor

During the COVID-19 pandemic, the availability of resources in hospitals around the world experiences significant fluctuations following successive infection waves. Thus, a clinically useful prediction tool needs to reckon with these dynamic scenarios for effectively assisting undertriage and overtriage decisions.

We developed and validated two subtypes of our predictor, CODOP-Ovt (from overtriage) and CODOP-Unt (from undertriage), intending to optimize the triage of patients at high risk of death upon arrival to the hospital and after their first blood analysis. CODOP-Ovt maximizes the negative predictive value or the detection of high-risk patients (high sensitivity) and it is meant for scenarios where overtriage is possible because hospital resources are not the main limitation. On the other side, CODOP-Unt maximizes the positive predictive value by trying to avoid the inclusion of false high-risk patients (high specificity) and it might be preferred in pandemic conditions when hospital resources are limited and undertriage needs to be considered.

Using the initial training cohort, CODOP-Ovt identified >95% of the patients that finally died in hospital at nine days before clinical resolution (Supplementary Figure 4A). As expected, this increase in sensitivity is concomitant with reduced specificity (60-70%; Supplementary Figure 4B). Notably, these metrics are within the range of recommended under- and overtriage levels ranging from 5-10% and 25-50%, respectively.^21^ The opposite results were obtained with CODOP-Unt, where more than 95% of the patients that survived were correctly predicted as low-risk (Supplementary Figure 4B) while 40-50% of the patients that died in hospital were not detected in advance (Supplementary Figure 4A). Confusion matrixes show similar overall performance for both CODOP subtypes in all test cohorts (Supplementary Tables 9-12).

Following, we constructed and evaluated an easy-to-use web-based application (https://gomezvarelalab.em.mpg.de/codop/) that offers the possibility to choose between CODOP-Ovt and CODOP-Unt. The web application includes a detailed description of the CODOP project and instructions on how to use the prediction tool. The web application has been tested using different devices, web browsers and operative systems (Supplementary Table 13). In all cases, predictions were calculated in less than 2 seconds for datasets up to 2 000 patients (data not shown). Further, the Data Protection Office of the Max Planck Society assisted in assuring the legal fit of the web application to the General Data Protection Regulation (GDPR).

To make a stringent external evaluation of this application with datasets collected from very different patient cohorts, we established a multinational collaboration with five hospitals from three Latin American countries (Figure 4A), which at the time of this evaluation were under a new surge of COVID-19 infections and admissions coinciding with the beginning of the Autumn-Winter season in the Southern Hemisphere. All these hospitals provided the values for the 12 features used by CODOP and measured in patients at the time of hospitalization between March 7th 2020 to June 7th 2021. Following, these data were uploaded to the two CODOP online subtypes and we obtained the mortality predictions that were compared to the real patient outcome (for which the online predictor was blinded).

**Figure 4.**
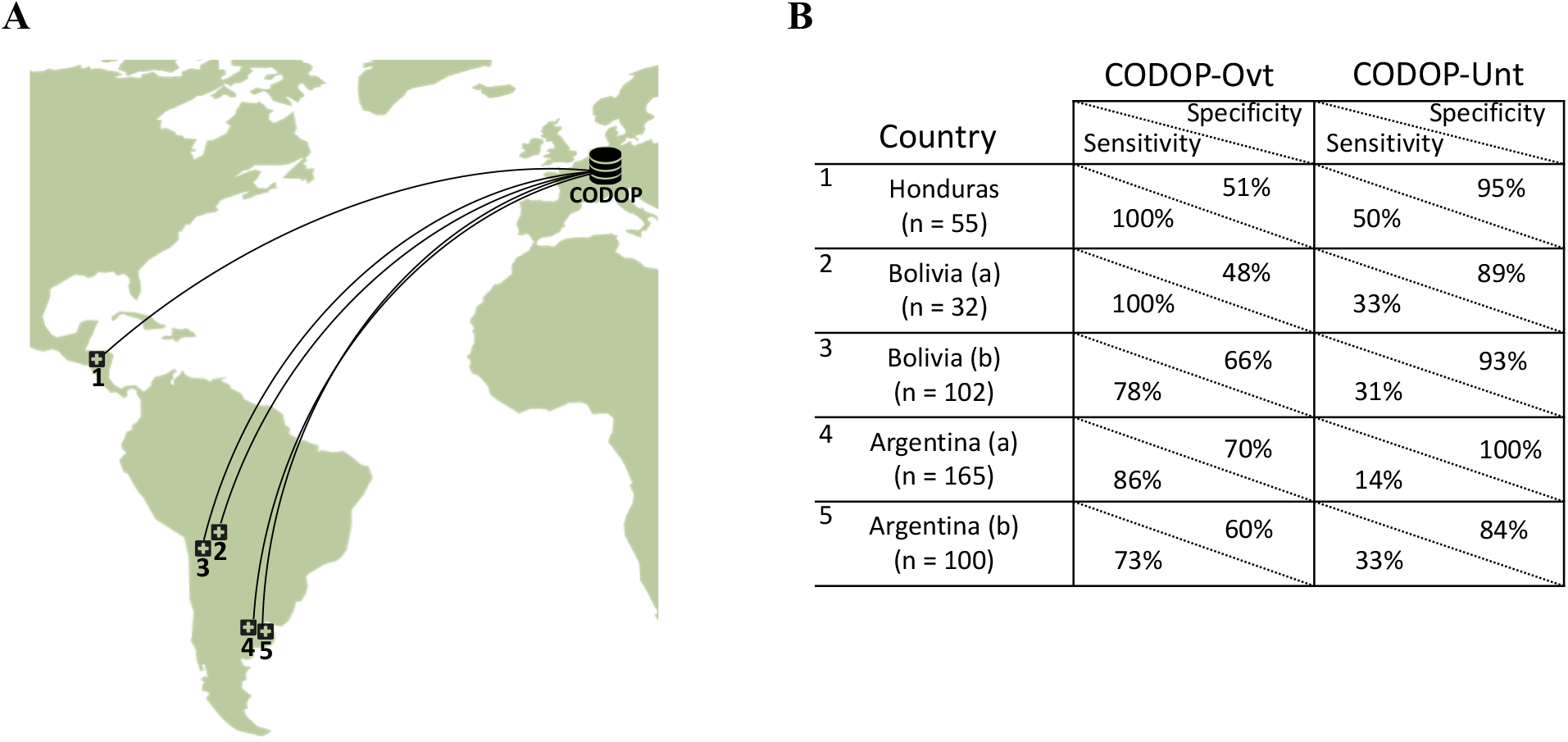
The geographical location of the external cohorts from 6 different Latin American hospitals used during the online evaluations (A) and performance of web calculators CODOP-Ovt and CODOP-Unt in these external cohorts (number of patients from each institution are indicated in parenthesis; B).

Importantly, AUROC values, horizon, calibration, and risk-stratification curves for CODOP-Ovt demonstrate the generalizability of the predictor (Supplementary Figure 5, Supplementary Table 5 and 6). A detailed analysis of the results indicates that if these were a prospective study, CODOP-Ovt would have identified the majority of the patients that finally died during hospitalization albeit wrongly classifying as high-risk a significant number of patients that finally survived (73-100% sensitivity and 48-70% specificity, respectively; Figure 4B and Supplementary Table 14). On the other side, the use of CODOP-Unt would have correctly triaged the vast majority of the survivors despite missing a significant number of patients that finally died (84-100% specificity and 14-50% sensitivity, respectively; Figure 4B and Supplementary Table 14). These results strongly suggest that the online version of CODOP could represent a useful clinical tool in the triage decision protocols.

## Discussion

The differential access and the low rates of COVID-19 vaccinations, the emergence of more contagious viral variants, and the waning of the immune protection project a longer period of health systems under severe strains leading to a bigger number of COVID-19 related deaths, particularly in resource-limited countries. A conflagration-like scenario will likely be the final set of the pandemic for many nations.^22^ As a result of an altruistic multicontinental effort, we developed and evaluated CODOP, a machine-learning-based online tool able to assist in triage decisions in hospitalized COVID-19 patients. CODOP uses 12 clinical parameters easy to collect in most hospitals. Its predictive performance among very different cohorts of patients strongly suggests its generalizability and supports its potential for improving patient care during this pandemic.

CODOP satisfies the Transparent Reporting of a multivariable prediction model for Individual Prognosis Or Diagnosis principles^23^ (TRIPOD; Supplementary Table 15), follows the recently proposed MINimum Information for Medical AI Reporting^24^ (MINIMAR; Supplementary Table 16 and 17), and it has been successfully checked for the risk of bias and applicability using the Prediction model study Risk of Bias Assessment Tool^25^ (PROBAST; Supplementary Table 18).

The use of such an early warning system like CODOP could potentially represent an important help in clinical decision-making including the prioritization of care and resource allocation. The novelty of the COVID-19 disease and its toll on the health systems has led to dozens of triage policies, many of them based on some form of Sequential Organ Failure Assessment (SOFA) scores.^26^ In addition, several machine learning-based prediction tools have been developed during this pandemic. However, independent validation studies have dismissed the clinical utility of all these models^3, 4^ and have indicated common pitfalls to be avoided such as small sample size, use of variables not easily measurable in most hospitals, lack of external evaluation datasets gathered in geographically different cohorts, etc. To avoid this “dataset shift” problem and aim to increase the generalization of CODOP, we set to satisfy the so-called stability property.^5^ For this we used an initial training and test cohorts encompassing 21 607 patients from more than 110 hospitals spread over Spain and the USA and gathered during three pandemic waves. Both the size, heterogeneity of the patient population (in terms of age range, ethnicity, comorbidities, etc.), and the myriad of clinical and analytical procedures performed during the pandemic, ensures a significant number of perturbations (shifts) in how the data were generated. This strategy seems to be supported by the stable performance of our predictor on the external online evaluation performed with five patient cohorts from three Latin American countries. We expect that future participation of more institutions from regions non-represented in our study (Africa, Asia) will improve the reproducibility and overall clinical utility of CODOP supporting subgroup-specific predictions (e. g., based on underlying comorbidities or ethnical background).

In addition to the characteristics of our cohorts, we hypothesized that the higher performance achieved by CODOP when compared to published mortality risk scores is due to the use of a group of biochemical parameters representing the main biological pathways involved in the pathogenesis of SARS-CoV-2. A very common clinical manifestation in critical COVID-19 patients is composed of a deregulated immune response and a robust inflammatory reaction (known as “hypercytokinemia” or “cytokine storm”), which ultimately leads to tissue injury.^27^ Recent reports show a downregulated type-I interferon response leading to an increase of neutrophils in severe COVID-19 patients.^28^. These findings go in line with our data showing alterations in several myeloid cells (eosinophils, monocytes) including an upregulation in the number of neutrophils (Supplementary Table 3). Myeloid cells are crucial for mounting a successful immune response against viruses and for the existence of hypercytokinemia.^29^ The increased level of CRP and LDH in our dataset and their predictive value could represent easy-to-measure hallmarks of the exacerbated inflammatory response associated with a high risk of COVID-19-related death. These and other model features linked to thromboembolic complications (i. e., D-dimer and Platelets) and organ failure (i. e., Creatinine), could represent a warning signature easy to evaluate at early stages of the infection, even before failure in major functions can be monitored.

The quality, availability, and consistency of biomedical data make reproducibility very challenging for machine learning tools applied to health^30^ (MLH). The reproducibility of MLH is of critical importance as predictions can affect human health care. Careful analysis indicates that CODOP fulfils the main performance criteria reached in other machine learning subfields when analysing the three main reproducibility principles. In comparison to previous studies, CODOP excels in the “Conceptual Reproducibility or Replicability” due to the use of geographically spread cohorts.^30^

The overall performance of CODOP has inherent limitations, some of them generalizable to any MLH. On the one side, the use of training and test datasets with a high degree of perturbations (see above) adds several sources of variability^31^: pre-analytical due to differences in blood sampling, analytical due to different laboratory protocols, intra- and inter-individual, and inter-hospital and geographical differences in clinical practices. As an additional factor, the high diversity of COVID-19 encompassing more than 60 disease subtypes^6^ sets a limitation in terms of the discriminability ability and the overall clinical utility of any MHL. In contrast to other predictors and to facilitate its use, CODOP does not take into account the level of care received by each patient (e.g., ICU versus basic care), which influences the outcome of the patient and perturbs the discrimination ability of CODOP (as predictions are made with the data from blood analyses at hospital admission). A clear example is a slightly lower performance of CODOP-Ovt (sensitivity of 73%) in the case of the “Hospital Vélez Sarsfield.. from Buenos Aires (named as Argentina (b) in Figure 4B), where all patients analysed by CODOP were finally treated in the ICU. On the other hand, CODOP-Unt would have correctly suggested triaging 84% of these patients already on the day of admission, therefore offering a significant clinical utility. Finally, the clinical utility of MHL has to take into account the changing pressure supported by hospitals during the successive pandemic waves. Our data support the strategy of using either CODOP-Unt or CODOP-Ovt as an effective first-line triage tool in the overall clinical decision procedure.

## Data Availability

The data that support the findings of this study are available on request from either the SEMI-COVID-19 Scientific Committee and the Registry Coordinating Centre or the different Latin American hospitals.

## Contributors

DGV conceived, planned and organize the study. DGV, RK and DP analysed the data. DGV and RK wrote the original draft of the manuscript. DGV, BL and NF conceived and set the online application. In addition to the authors mentioned above, the rest of the authors were responsible for performing the different steps necessary for the collection of the blood datasets used in this study. All authors contributed to reading and approving the final version of the manuscript. The corresponding author (DGV) attests that all listed authors meet authorship criteria and that no others meeting the criteria have been omitted.

## Acknowledgements

We gratefully acknowledge all the investigators and staff from the SEMI-COVID-19 Registry and from the five Latin American hospitals who participate in the collection of the patient data (see Appendix 1).

We also gratefully thank the Data Protection Office of the Max Planck Society assisted for assuring the legal fit of the web application to the General Data Protection Regulation (GDPR), and to the IT team of the Max Planck Institute of Experimental Medicine for their support in setting up and maintaining the CODOP web calculator.

