## Supplementary Figures and Tables for "Development and evaluation of a machine learning-based in-hospital COvid-19 Disease Outcome Predictor (CODOP): a multicontinental retrospective study"

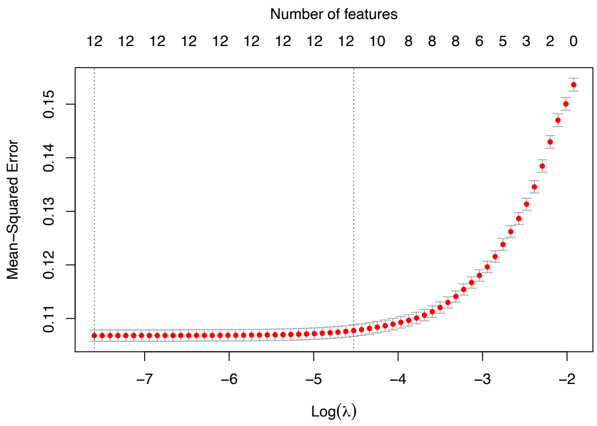

**Supplementary Figure 1.** Optimisation of the final COPOD model by selecting predictors using the least absolute shrinkage and selection operator (LASSO) method. The mean squared error is plotted versus log of the Penalty parameter (λ). Figure is produced by function cv.glmnet from R package glmnet.

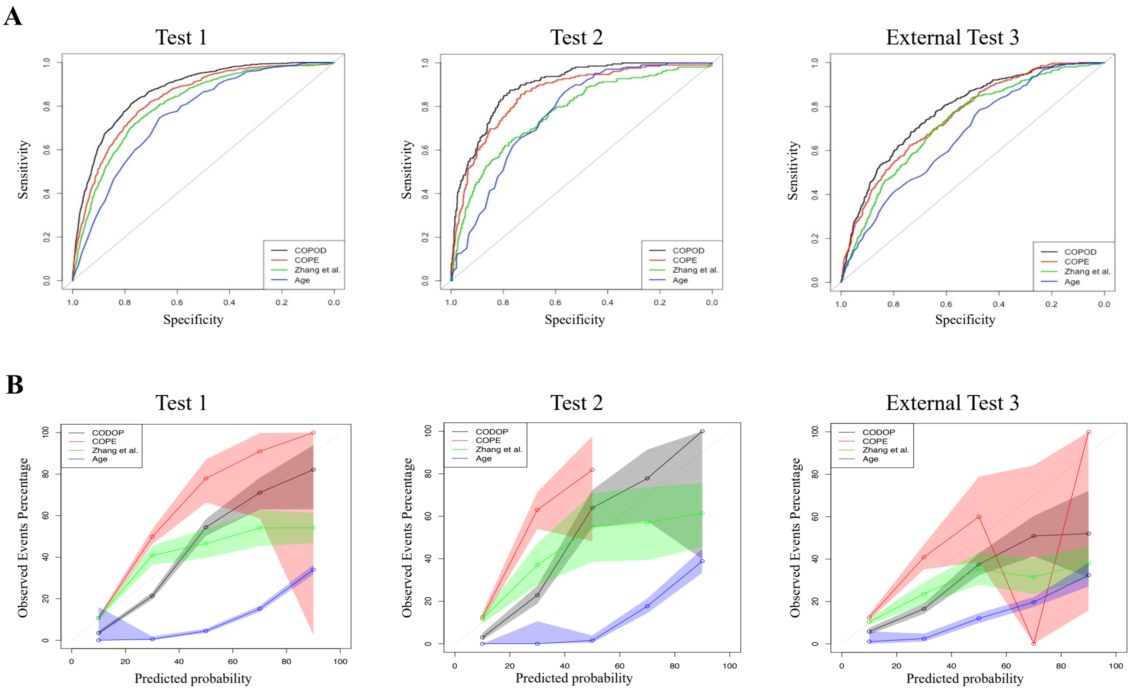

**Supplementary Figure 2.** Discriminatory ability (using area under receiver operating curves or AUROC; A) and calibration curves (B) for CODOP, COPE, Zhang et al., and Age in the test datasets.

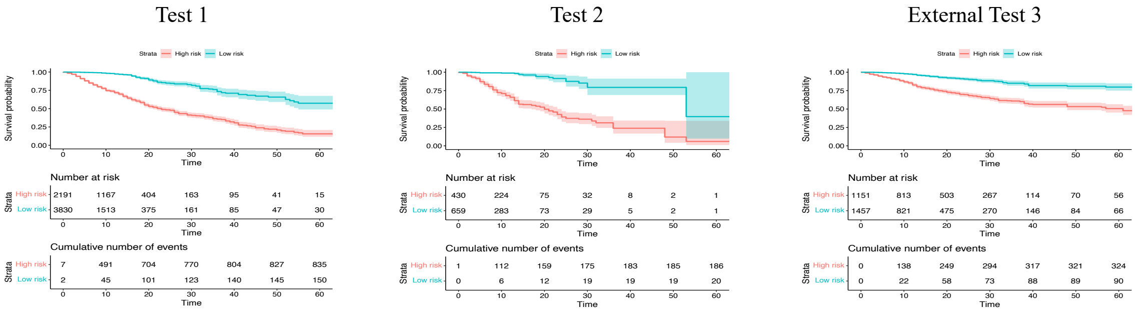

**Supplementary Figure 3.** Survival analysis of CODOP in the test datasets.

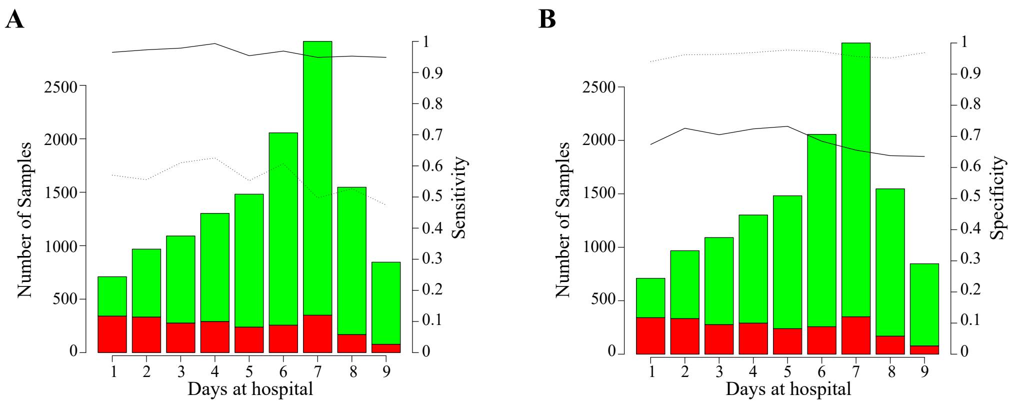

**Supplementary Figure 4.** Horizon analysis in the training dataset for sensitivity (A) and specificity (B). The black solid line is CODOP-Ovt and black dotted line is CODOP-Unt.

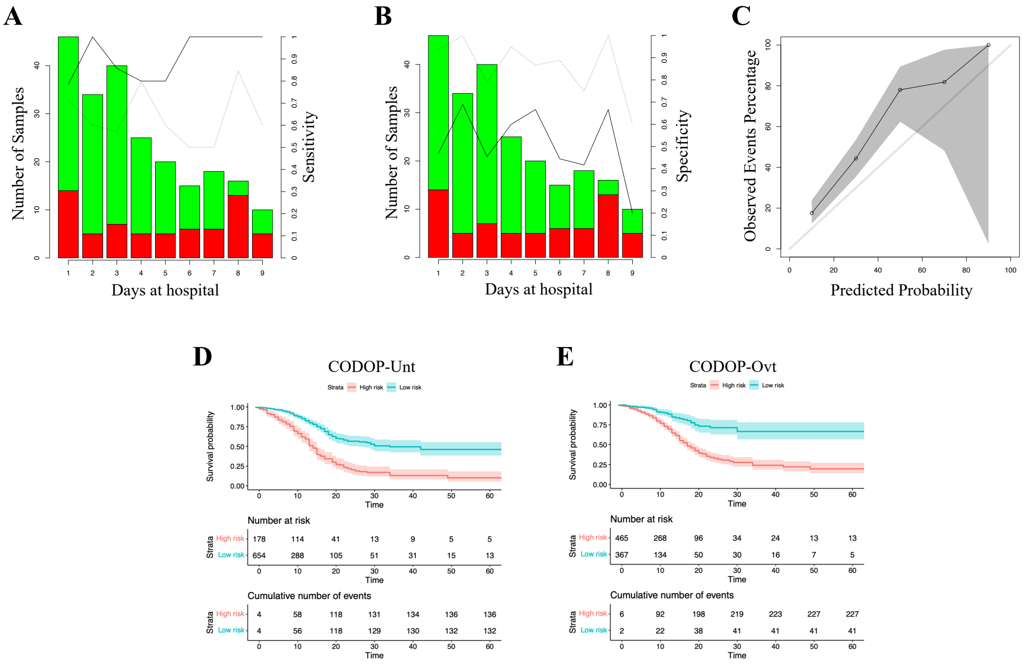

**Supplementary Figure 5.** Horizon analysis for all the Latin American external datasets together for sensitivity (A) and specificity (B) for CODOP-Unt (black dotted line) and CODOP-Ovt (black solid line). Calibration plot for all the Latin American external datasets together (C). Survival analyses for all the Latin American external datasets together for CODOP-Unt (D) and CODOP-Ovt (E). In horizon analysis, black dotted line is CODOP-Ovt and the black solid line is CODOP-Unt.

**Supplementary Table 1**. Features used during CODOP development with the training cohort, the values used for imputation, and the percentage of missing values. Numerical variables are reported by median (Md) and interquartile range (IQR).

| Variable | Imputed value | Md (IQR) | Missing % |
|---|---|---|---|
| Age (years) | 66·67911 | 68 (56-79) | 0·0 |
| Sex (male, female) | none | 6 775 females and 9 127 males | 0·0 |
| Hemoglobin (g/dL) | 13·33201 | 13 (12-15) | 1·7 |
| Platelet Count (x 10^6/L) | 250 097·7 | 223 000 (164 000-311 000) | 1·8 |
| Eosinophils (x 10^6/L) | 63·81817 | 10 (0-100) | 3·0 |
| Lymphocytes (x 10^6/L) | 1 243·575 | 1 000 (700-1 420) | 1·9 |
| Neutrophils (x 10^6/L) | 5 525·894 | 4 490 (3 090-6 800) | 2·2 |
| Monocytes (x 10^6/L) | 535·8804 | 470 (300-660) | 2·7 |
| C-Reactive Protein (mg/L) | 74·48964 | 41 (12-108) | 4·6 |
| Creatinine (mg/dL) | 1·156574 | 1 (1-1) | 2·0 |
| Lactate Dehydrogenase (U/L) | 363·9083 | 306 (234-424) | 13·0 |
| Aspartate aminotransferase (U/L) | 49·27098 | 35 (24-53) | 18·4 |
| Alanine aminotransferase (U/L) | 48·99699 | 32 (20-54 | 7·4 |
| Total bilirrubin (mg/dL) | 0·6429202 | 1 (0-1) | 26·5 |
| Serum Sodium (mmol/L) | 138·4268 | 138 (136-141) | 2·6 |
| Serum Potassium (mmol/L) | 4·178441 | 4 (4-4) | 3·7 |
| Glucose (mg/dL) | 124·2852 | 108 (92-135) | 5·2 |
| Prothrombin time (s) | 19·99798 | 13 (12-14) | 35·8 |
| Fibrinogen (mg/dL) | 608·0043 | 601 (497-713) | 37·0 |
| Dimer (ng/mL) | 2 122·158 | 672 (370-1 320) | 21·7 |

**Supplementary Table 2.** Feature occurrence during CODOP development iterative step of 100 iterations, and final list of selected features.

| Variable | Occurrences | Selected |
|---|---|---|
| Age (years) | 100 | X |
| Sex (male=0, female=1) | 95 | |
| Hemoglobin (g/dL) | 95 | |
| Platelet Count (x 10^6/L) | 100 | X |
| Eosinophils (x 10^6/L) | 100 | X |
| Lymphocytes (x 10^6/L) | 0 | |
| Neutrophils (x 10^6/L) | 100 | X |
| Monocytes (x 10^6/L) | 100 | X |
| C-Reactive Protein (mg/L) | 100 | X |
| Creatinine (mg/dL) | 100 | X |
| Lactate Dehydrogenase (U/L) | 100 | X |
| Aspartate aminotransferase (U/L) | 0 | |
| Alanine aminotransferase (U/L) | 55 | |
| Total bilirrubin (mg/dL) | 0 | |
| Serum Sodium (mmol/L) | 100 | X |
| Serum Potassium (mmol/L) | 100 | X |
| Glucose (mg/dL) | 100 | X |
| Prothrombin time (s) | 0 | |
| Fibrinogen (mg/dL) | 0 | |
| Dimer (ng/mL) | 100 | X |

**Supplementary Table 3.** Descriptive statistics of training dataset by survival status. Variables are reported by median and interquartile range.

| Variable | Non-survival | Survival | P-value |
|---|---|---|---|
| Age (years) | 80 (73-87) | 65 (53-75) | <0·001 |
| Platelet Count (x 10^6/L) | 189 000 (142 000-255 000) | 232 000 (170 000-323 000) | <0·001 |
| Neutrophils(x 10^6/L) | 6 700 (4260-10 100) | 4 180 (2 920-6 100) | <0·001 |
| C-Reactive Protein (mg/L) | 114 (43-201) | 32 (10-86) | <0·001 |
| Creatinine (mg/dL) | 1 (1-2) | 1 (1-1) | <0·001 |
| Lactate Dehydrogenase (U/L) | 449 (325-635) | 287 (225-384) | <0·001 |
| Serum Sodium (mmol/L) | 139 (135-143) | 138 (136-140) | <0·001 |
| Serum Potassium (mmol/L) | 4 (4-5) | 4 (4-4) | <0·001 |
| Glucose (mg/dL) | 131 (107-175) | 104 (90-126) | <0·001 |
| D-Dimer (ng/mL) | 1 282 (653-2 816·5) | 601 (342-1110) | <0·001 |
| Eosinophils (x 10^6/L) | 0 (0-20) | 10 (0-100) | <0·001 |
| Monocytes (x 10^6/L) | 400 (240-600) | 490 (310-680) | <0·001 |

**Supplementary Table 4.** Univariable analysis. For all variables a univariable model was evaluated in training dataset. The best univariable model was selected based on average ranking of AUROC, accuracy, sensitivity and specificity. The best variable was age (average ranking 3·50) followed by creatinine (7·50) and D-Dimer (7·75).

| Variable | AUROC | Accuracy | Sensitivity | Specificity | Rank |
|---|---|---|---|---|---|
| Age (years) | 78·9 | 70·1 | 77·3 | 68·4 | 3·50 |
| Hemoglobin (g/dL) | 57·2 | 65·9 | 59·9 | 28·1 | 8·50 |
| Platelet Count (x 10^6/L) | 62·3 | 50·8 | 25·3 | 56·7 | 14·50 |
| Eosinophils (x 10^6/L) | 62·5 | 51·1 | 24·3 | 54·6 | 12·00 |
| Lymphocytes (x 10^6/L) | 70·5 | 67·7 | 37·7 | 31·0 | 11·00 |
| Neutrophils (x 10^6/L) | 69·7 | 66·8 | 63·7 | 67·5 | 12·50 |
| Monocytes (x 10^6/L) | 58·1 | 65·4 | 56·1 | 29·6 | 8·75 |
| C-Reactive Protein (mg/L) | 72·6 | 67·3 | 68·0 | 67·2 | 10·50 |
| Creatinine (mg/dL) | 63·6 | 81·1 | 34·6 | 92·0 | 7·50 |
| Lactate Dehydrogenase (U/L) | 73·1 | 64·2 | 74·9 | 61·7 | 11·50 |
| Aspartate aminotransferase (U/L) | 59·7 | 50·2 | 70·8 | 44·8 | 9·25 |
| Alanine aminotransferase (U/L) | 55·0 | 61·1 | 19·7 | 70·7 | 8·50 |
| Total bilirrubin (mg/dL) | 51·9 | 57·1 | 69·2 | 36·8 | 14·75 |
| Serum Sodium (mmol/L) | 53·7 | 78·0 | 24·5 | 90·5 | 15·75 |
| Serum Potassium (mmol/L) | 55·1 | 67·9 | 36·0 | 75·3 | 11·25 |
| Glucose (mg/dL) | 69·3 | 64·1 | 67·3 | 63·4 | 13·50 |
| Prothrombin time (s) | 56·4 | 52·7 | 71·3 | 41·7 | 8·75 |
| Fibrinogen (mg/dL) | 54·2 | 59·1 | 74·8 | 32·9 | 10·00 |
| D-Dimer (ng/mL) | 68·3 | 61·5 | 70·2 | 59·4 | 7·75 |

**Supplementary Table 5.** Evaluation metrics sensitivity, specificity, accuracy and area under receiver operating curve (AUROC) in all datasets.

| | Sensitivity | Specificity | Accuracy | AUROC |
|---|---|---|---|---|
| **Training dataset** | | | | |
| CODOP | 0·839 | 0·784 | 0·794 | 0·889 (0·885-0·894) |
| COPE | 0·791 | 0·787 | 0·787 | 0·864 (0·859-0·869) |
| Zhang et· al· | 0·796 | 0·701 | 0·719 | 0·816 (0·811-0·822) |
| Univariable | 0·773 | 0·684 | 0·701 | 0·789 (0·783-0·795) |
| **Test 1** | | | | |
| CODOP | 0·843 | 0·731 | 0·750 | 0·869 (0·856-0·881) |
| COPE | 0·765 | 0·755 | 0·757 | 0·834 (0·821-0·847) |
| Zhang et· al· | 0·787 | 0·686 | 0·703 | 0·807 (0·793-0·821) |
| Univariable | 0·770 | 0·625 | 0·649 | 0·758 (0·743-0·773) |
| **Test 2** | | | | |
| CODOP | 0·899 | 0·724 | 0·758 | 0·893 (0·871-0·914) |
| COPE | 0·788 | 0·772 | 0·775 | 0·857 (0·829-0·885) |
| Zhang et· al· | 0·784 | 0·608 | 0·642 | 0·772 (0·735-0·808) |
| Univariable | 0·837 | 0·587 | 0·635 | 0·768 (0·738-0·799) |
| **External Test 3** | | | | |
| CODOP | 0·748 | 0·691 | 0·705 | 0·780 (0·758-0·803) |
| COPE | 0·528 | 0·735 | 0·671 | 0·754 (0·730-0·778) |
| Zhang et· al· | 0·676 | 0·642 | 0·652 | 0·724 (0·699-0·750) |
| Univariable | 0·290 | 0·799 | 0·643 | 0·697 (0·644-0·697) |
| **Online External** | | | | |
| CODOP-Unt | 0·319 | 0·956 | 0·746 | 0·774 (0·724-0·823) |
| CODOP-Ovt | 0·768 | 0·638 | 0·679 | 0·774 (0·724-0·823) |

**Supplementary Table 6.** Statistics for calibration plots. Intercept and slope are for linear fit and root mean squared error (RMSE) is calculated between the fitted line and the diagonal line.

| | **Intercept** | **Slope** | **RMSE** |
|---|---|---|---|
| **Training dataset** | | | |
| CODOP | -0·473 | 1·051 | 0·0100 |
| COPE | 25·801 | 0·739 | 0·0490 |
| Zhang et· al· | 15·594 | 0·600 | 0·0193 |
| Age | -12·971 | 0·531 | 0·1556 |
| **Test 1** | | | |
| CODOP | -5·257 | 1·035 | 0·0040 |
| COPE | 11·015 | 1·098 | 0·0343 |
| Zhang et· al· | 16·253 | 0·500 | 0·0334 |
| Age | -9·829 | 0·413 | 0·1838 |
| **Test 2** | | | |
| CODOP | -8·716 | 1·245 | 0·0091 |
| COPE | 0·4624 | 1·733 | 0·0702 |
| Zhang et· al· | 14·591 | 0·597 | 0·0212 |
| Age | -12·233 | 0·477 | 0·1740 |
| **External Test 3** | | | |
| CODOP | 0·865 | 0·633 | 0·0433 |
| COPE | 9·159 | 0·670 | 0·1045 |
| Zhang et· al· | 11·922 | 0·316 | 0·0889 |
| Age | -6·423 | 0·399 | 0·1626 |
| **Online External** | | | |
| CODOP-Unt and CODOP-Ovt | 13·786 | 1·0115 | 0·0258 |

**Supplementary Table 7.** Ratio on deaths in different datasets.

| Dataset | Number of deaths | Percentage of deaths |
|---|---|---|
| Training dataset | 3147 | 19·8 |
| Test 1 | 530 | 17·0 |
| Test 2 | 108 | 19·1 |
| External Test 3 | 331 | 16·4 |
| Online External | 138 | 30·0 |

**Supplementary Table 8.** Area under receiver operating curve and 95% confidence interval in horizon analysis using the training cohort.

| Days at hospital | CODOP | COPE | Zhang et· al· | Univariable |
|---|---|---|---|---|
| 1 | 0·910 (0·889-0·931) | 0·907 (0·885-0·929) | 0·858 (0·830-0·886) | 0·845 (0·816-0·874) |
| 2 | 0·939 (0·924-0·954) | 0·931 (0·915-0·947) | 0·874 (0·850-0·897) | 0·863 (0·839-0·887) |
| 3 | 0·950 (0·938-0·963) | 0·929 (0·914-0·944) | 0·894 (0·872-0·916) | 0·813 (0·787-0·840) |
| 4 | 0·960 (0·950-0·970) | 0·930 (0·913-0·946) | 0·913 (0·894-0·932) | 0·835 (0·811-0·858) |
| 5 | 0·947 (0·934-0·960) | 0·911 (0·891-0·932) | 0·919 (0·902-0·937) | 0·824 (0·798-0·850) |
| 6 | 0·943 (0·930-0·955) | 0·910 (0·893-0·926) | 0·898 (0·878-0·919) | 0·796 (0·771-0·821) |
| 7 | 0·905 (0·888-0·921) | 0·852 (0·832-0·873) | 0·876 (0·857-0·894) | 0·736 (0·710-0·762) |
| 8 | 0·910 (0·888-0·931) | 0·851 (0·820-0·882) | 0·886 (0·865-0·907) | 0·764 (0·731-0·797) |
| 9 | 0·910 (0·879-0·942) | 0·845 (0·800-0·889) | 0·828 (0·775-0·882) | 0·722 (0·667-0·777) |

**Supplementary Table 9.** Confusion matrix in Training dataset.

| CODOP-Unt | Truth | |
|---|---|---|
| Predicted | Survived | Died |
| Survived | 23 645 | 2 988 |
| Died | 1 308 | 2 852 |

| CODOP-Ovt | Truth | |
|---|---|---|
| Predicted | Survived | Died |
| Survived | 14 845 | 292 |
| Died | 10 108 | 5 548 |

**Supplementary Table 10.** Confusion matrix in Test 1 dataset.

| CODOP-Unt | Truth | |
|---|---|---|
| Predicted | Survived | Died |
| Survived | 4 669 | 504 |
| Died | 357 | 491 |

| CODOP-Ovt | Truth | |
|---|---|---|
| Predicted | Survived | Died |
| Survived | 2 648 | 53 |
| Died | 2 378 | 942 |

**Supplementary Table 11.** Confusion matrix in Test 2 dataset.

| CODOP-Unt | Truth | |
|---|---|---|
| Predicted | Survived | Died |
| Survived | 827 | 100 |
| Died | 54 | 108 |

| CODOP-Ovt | Truth | |
|---|---|---|
| Predicted | Survived | Died |
| Survived | 442 | 4 |
| Died | 439 | 204 |

**Supplementary Table 12.** Confusion matrix in External Test 3.

| **CODOP-Unt** | **Truth** | |
|---|---|---|
| Predicted | Survived | Died |
| Survived | 1 872 | 200 |
| Died | 317 | 219 |

| **CODOP-Ovt** | **Truth** | |
|---|---|---|
| Predicted | Survived | Died |
| Survived | 929 | 34 |
| Died | 1 260 | 385 |

**Supplementary Table 13.** Devices, web browsers and operative systems used for testing the CODOP web calculator

| **Device model** | **Operative system** | **Web browsers** |
|---|---|---|
| Dell L5510 notebook | Ubuntu 20.04 LTS | Firefox (v86.0.1 |
| Dell L5510 notebook | Ubuntu 20.04 LTS | Chromium (v 89.0.4339.90) |
| MacBook Pro | High Sierra (v10.13.6) | Safari (v13.1.2) |
| MacBook Pro | High Sierra (v10.13.6) | Firefox (v78.8.0) |
| Pixel 5 Phone | Android 11 Phone | Firefox |
| iPhone SE | 14.4 | Safari |

**Supplementary Table 14.** Confusion matrix in the Online External evaluation dataset.

**All hospitals**

| | | |
|---|---|---|
| **CODOP-Unt** | **Truth** | |
| Predicted | Survived | Died |
| Survived | 268 | 94 |
| Died | 12 | 44 |

| **CODOP-Ovt** | **Truth** | |
|---|---|---|
| Predicted | Survived | Died |
| Survived | 178 | 32 |
| Died | 102 | 106 |

**Honduras**

| **CODOP-Unt** | **Truth** | |
|---|---|---|
| Predicted | Survived | Died |
| Survived | 39 | 2 |
| Died | 2 | 2 |

| **CODOP-Ovt** | **Truth** | |
|---|---|---|
| Predicted | Survived | Died |
| Survived | 21 | 0 |
| Died | 20 | 4 |

**Argentina (a)**

| **CODOP-Unt** | **Truth** | |
|---|---|---|
| Predicted | Survived | Died |
| Survived | 143 | 6 |
| Died | 0 | 1 |

| **CODOP-Ovt** | **Truth** | |
|---|---|---|
| Predicted | Survived | Died |
| Survived | 100 | 1 |
| Died | 43 | 6 |

**Bolivia (a)**

| **CODOP-Unt** | **Truth** | |
|---|---|---|
| Predicted | Survived | Died |
| Survived | 24 | 2 |
| Died | 3 | 1 |

| CODOP-Ovt | Truth | |
|---|---|---|
| Predicted | Survived | Died |
| Survived | 13 | 0 |
| Died | 14 | 3 |

### Bolivia (b)

| CODOP-Unt | Truth | |
|---|---|---|
| Predicted | Survived | Died |
| Survived | 41 | 34 |
| Died | 3 | 15 |

| CODOP-Ovt | Truth | |
|---|---|---|
| Predicted | Survived | Died |
| Survived | 29 | 11 |
| Died | 15 | 38 |

### Argentina (b)

| CODOP-Unt | Truth | |
|---|---|---|
| Predicted | Survived | Died |
| Survived | 21 | 50 |
| Died | 4 | 25 |

| CODOP-Ovt | Truth | |
|---|---|---|
| Predicted | Survived | Died |
| Survived | 15 | 20 |
| Died | 10 | 55 |

**Supplementary Table 15.** TRIPOD Checklist; Prediction Model Development

| Section/Topic | Item | Checklist Item | Page |
|---|---|---|---|
| **Title and abstract** | | | |
| Title | 1 | Identify the study as developing and/or validating a multivariable prediction model, the target population, and the outcome to be predicted. | 1 |
| Abstract | 2 | Provide a summary of objectives, study design, setting, participants, sample size, predictors, outcome, statistical analysis, results, and conclusions. | 1 |
| **Introduction** | | | |
| Background and objectives | 3a | Explain the medical context (including whether diagnostic or prognostic) and rationale for developing or validating the multivariable prediction model, including references to existing models. | 2-4 |
| | 3b | Specify the objectives, including whether the study describes the development or validation of the model or both. | 2-3 |
| **Methods** | | | |
| Source of data | 4a | Describe the study design or source of data (e.g., randomized trial, cohort, or registry data), separately for the development and validation data sets, if applicable. | 11 |
| | 4b | Specify the key study dates, including start of accrual; end of accrual; and, if applicable, end of follow-up. | 11 |
| Participants | 5a | Specify key elements of the study setting (e.g., primary care, secondary care, general population) including number and location of centres. | 11 |
| | 5b | Describe eligibility criteria for participants. | 11 |
| | 5c | Give details of treatments received, if relevant. | n/a |
| Outcome | 6a | Clearly define the outcome that is predicted by the prediction model, including how and when assessed. | 11 |
| | 6b | Report any actions to blind assessment of the outcome to be predicted. | 7, 13 |
| Predictors | 7a | Clearly define all predictors used in developing or validating the multivariable prediction model, including how and when they were measured. | 11-12 |
| | 7b | Report any actions to blind assessment of predictors for the outcome and other predictors. | 7, 13 |
| Sample size | 8 | Explain how the study size was arrived at. | 11 |
| Missing data | 9 | Describe how missing data were handled (e.g., complete-case analysis, single imputation, multiple imputation) with details of any imputation method. | 4,11 |
| Statistical analysis methods | 10a | Describe how predictors were handled in the analyses. | 11 |
| | 10b | Specify type of model, all model-building procedures (including any predictor selection), and method for internal validation. | 12 |
| | 10d | Specify all measures used to assess model performance and, if relevant, to compare multiple models. | 12-13 |
| Risk groups | 11 | Provide details on how risk groups were created, if done. | 12 |
| **Results** | | | |
| Participants | 13a | Describe the flow of participants through the study, including the number of participants with and without the outcome and, if applicable, a summary of the follow-up time. A diagram may be helpful. | 11 Fig. 1 |
| | 13b | Describe the characteristics of the participants (basic demographics, clinical features, available predictors), including the number of participants with missing data for predictors and outcome. | Suppl. Table 1 and Suppl. Table 16 |
| Model development | 14a | Specify the number of participants and outcome events in each analysis. | Figure1 and page 11 |
| | 14b | If done, report the unadjusted association between each candidate predictor and outcome. | Suppl. Table 4 |
| Model specification | 15a | Present the full prediction model to allow predictions for individuals (i.e., all regression coefficients, and model intercept or baseline survival at a given time point). | Suppl. Table 15 |
| | 15b | Explain how to the use the prediction model. | 6 |
| Model performance | 16 | Report performance measures (with CIs) for the prediction model. | Fig. 2-4 Suppl, Table 5 & 7 |
| **Discussion** | | | |
| Limitations | 18 | Discuss any limitations of the study (such as nonrepresentative sample, few events per predictor, missing data). | 9-10 |
| Interpretation | 19b | Give an overall interpretation of the results, considering objectives, limitations, and results from similar studies, and other relevant evidence. | 9-10 |
| Implications | 20 | Discuss the potential clinical use of the model and implications for future research. | 10 |
| **Other information** | | | |
| Supplementary information | 21 | Provide information about the availability of supplementary resources, such as study protocol, Web calculator, and data sets. | 6 Suppl. |

| | | | |
|---|---|---|---|
| | | | Tables 2, 3, 6, 8-13 Suppl. Fig. 1-5 |
| Funding | 22 | Give the source of funding and the role of the funders for the present study. | 13 |

**Supplementary Table 16.** CODOP prediction model

| Variable | Coefficient | Example values |
|---|---|---|
| Intercept | $A_0$=-1·330923e+00 | - |
| Age | $A_1$=6·004188e-03 | $x_1$=73 |
| Platelet Count | $A_2$=-2·472077e-07 | $x_2$=223 000 |
| Eosinophils | $A_3$=-9·174623e-06 | $x_3$=12 |
| Neutrophils | $A_4$=7·551235e-06 | $x_4$=4 490 |
| Monocytes | $A_5$=-7·367404e-06 | $x_5$=470 |
| C-Reactive Protein | $A_6$=7·458099e-04 | $x_6$=41 |
| Creatinine | $A_7$=3·664203e-02 | $x_7$=1 |
| Lactate Dehydrogenase | $A_8$=2·722994e-04 | $x_8$=326 |
| Serum Sodium | $A_9$=5·894423e-03 | $x_9$=138 |
| Serum Potassium | $A_{10}$=1·236685e-02 | $x_{10}$=4 |
| Glucose | $A_{11}$=6·397398e-04 | $x_{11}$=108 |
| D-Dimer | $A_{12}$=3·290633e-07 | $x_{12}$=1 500 |
| Model | | |
| $C=A_0+A_1*x_1+A_2*x_2+A_3*x_3+A_4*x_4+A_5*x_5+A_6*x_6+A_7*x_7+A_8*x_8+A_9*x_9+A_{10}*x_{10}+A_{11}*x_{11}+A_{12}*x_{12}$ | | |
| COPOD-Ovt: If C<0·1677094, then 0. Otherwise 1. | | |
| COPOD-Unt: If C<0·3901752, then 0. Otherwise 1. | | |
| Example | | |
| -1·330923+0·006004188*68+…+0·0000003290633*1500=0·1710605 | | |
| COPOD-Ovt=1 and COPOD-Unt=0 | | |

**Supplementary Table 17.** Race and ethnicity

#### SEMI Cohorts

| **Race** | **Caucasic** | **African-American** | **Hispanic** | **Asian** | **Other** |
|---|---|---|---|---|---|
| Training cohort | 14 087 | 69 | 1 302 | 71 | 152 |
| Test 1 cohort | 2 657 | 39 | 293 | 23 | 79 |
| Test 2 cohort | 514 | 2 | 30 | 0 | 10 |

#### External Test 3 Cohort (Del Valle etal)

| | |
|---|---|
| African-American | 391 |
| American Indian or Alaskan | 1 |
| Asian Indian | 30 |
| Bangladeshi | 18 |
| Chinese | 22 |
| Congolese | 1 |
| Filipino | 15 |
| Ghanaian | 1 |
| Haitian | 7 |
| Hispanic | 881 |
| Indonesian | 3 |
| Jamaican | 4 |
| Japanese | 3 |
| Korean | 4 |
| Nepalese | 1 |
| Nigerian | 2 |
| Pakistani | 2 |
| Samoan | 1 |
| Trinidadian | 3 |
| Ugandan | 2 |
| West indian | 4 |
| White | 405 |
| Other | 220 |

#### Online External Cohort (Latin America Cohort)

| Asian | Caucasian | Hispanic |
|---|---|---|
| 3 | 45 | 370 |

**Supplementary Table 18.**

PROBAST (Prediction model study Risk Of Bias Assessment Tool)

Published in Annals of Internal Medicine (freely available):

1. PROBAST: A Tool to Assess the Risk of Bias and Applicability of Prediction Model Studies
2. PROBAST: A Tool to Assess Risk of Bias and Applicability of Prediction Model Studies: Explanation and Elaboration

**What does PROBAST assess?**
PROBAST assesses both the *risk of bias* and *concerns regarding applicability* of a study that evaluates (develops, validates or updates) a multivariable diagnostic or prognostic prediction model. It is designed to assess primary studies included in a systematic review.

*Bias* occurs if systematic flaws or limitations in the design, conduct or analysis of a primary study distort the results. For the purpose of prediction modelling studies, we have defined *risk of bias* to occur when shortcomings in the study design, conduct or analysis lead to systematically distorted estimates of a model's predictive performance or to an inadequate model to address the research question. Model predictive performance is typically evaluated using calibration, discrimination and sometimes classification measures, and these are likely inaccurately estimated in studies with high risk of bias. *Applicability* refers to the extent to which the prediction model from the primary study matches your systematic review question, for example in terms of the participants, predictors or outcome of interest.

A primary study may include the development and/or validation or update of more than one prediction model. A PROBAST assessment should be completed for each distinct model that is developed, validated or updated (extended) for making individualised predictions. Where a publication assesses multiple prediction models, only complete a PROBAST assessment for those models that meet the inclusion criteria for your systematic review. Please note that subsequent use of the term "model" includes derivatives of models, such as simplified risk scores, nomograms, or recalibrations of models.

PROBAST is not designed for all multivariable diagnostic or prognostic studies. For example, studies using multivariable models to identify predictors associated with an outcome but not attempting to develop a model for making individualised predictions are not covered by PROBAST.

PROBAST includes four steps.

| Step | Task | When to complete |
|---|---|---|
| **1** | Specify your systematic review question(s) | Once per systematic review |
| **2** | Classify the type of prediction model evaluation | Once for each model of interest in each publication being assessed, for each relevant outcome |
| **3** | Assess risk of bias and applicability | Once for each development and validation of each distinct prediction model in a publication |
| **4** | Overall judgment | Once for each development and validation of each distinct prediction model in a publication |

If this is your first time using PROBAST, we strongly recommend reading the detailed explanation and elaboration (E&E, see link above) paper and to check the examples on www.probast.org

**Step 1: Specify your systematic review question**

State your systematic review question to facilitate the assessment of the applicability of the evaluated models to your question. *The following table should be completed once per systematic review.*

| Criteria | Specify your systematic review question |
|---|---|
| *Intended use of model:* | Clinical outcome prediction (in hospital death or survival) |
| ***Participants** including selection criteria and setting:* | All subjects diagnosed with COVID-19 within select hospitals |
| ***Predictors** (used in prediction modelling), including types of predictors (e.g. history, clinical examination, biochemical markers, imaging tests), time of measurement, specific measurement issues (e.g., any requirements/ prohibitions for specialized equipment):* | Age (years), Sex (male/female), Days at hospital, Hemoglobin (g/dL), Platelet Count (x 10^6/L), Eosinophils (x 10^6/L), Lymphocytes (x 10^6/L), Neutrophils (x 10^6/L), Monocytes (x 10^6/L), C-Reactive Protein (mg/L), Creatinine (mg/dL), Lactate Dehydrogenase (U/L), Aspartate aminotransferase (U/L), Alanine aminotransferase (U/L), Total bilirrubin (mg/dL), Serum Sodium (mmol/L), Serum Potassium (mmol/L), Glucose (mg/dL), Prothrombin time (s), Fibrinogen (mg/dL) and Dimer (ng/mL). From the 5$^{th}$ February 2020 to the 7$^{th}$ June 2021. |
| *Outcome to be predicted:* | Binary outcome Survive/Death |

**Step 2: Classify the type of prediction model evaluation**

Use the following table to classify the evaluation as model development, model validation or model update, or combination. Different signalling questions apply for different types of prediction model evaluation. If the evaluation does not fit one of these classifications then PROBAST should not be used.

| **Classify the evaluation based on its aim** | | | |
|---|---|---|---|
| **Type of prediction study** | **PROBAST boxes to complete** | **Tick as appropriate** | **Definition for type of prediction model study** |
| Development only | Development | | Prediction model development without external validation. These studies may include internal validation methods, such as bootstrapping and cross-validation techniques. |
| Development and validation | Development and validation | X | Prediction model development combined with external validation in other participants in the same article. |
| Validation only | Validation | | External validation of existing (previously developed) model in other participants. |

*This table should be completed once for each publication being assessed and for each relevant outcome in your review.*

| |
|---|
| **Publication reference** |
| **Models of interest** |
| **Outcome of interest** |

**Step 3: Assess risk of bias and applicability**

PROBAST is structured as four key domains. Each domain is judged for risk of bias (low, high or unclear) and includes signalling questions to help make judgements. Signalling questions are rated as yes (Y), probably yes (PY), probably no (PN), no (N) or no information (NI). All signalling questions are phrased so that "yes" indicates absence of bias. Any signalling question rated as "no" or "probably no" flags the potential for bias; you will need to use your judgement to determine whether the domain should be rated as "high", "low" or "unclear" risk of bias. The guidance document contains further instructions and examples on rating signalling questions and risk of bias for each domain.

The first three domains are also rated for concerns regarding applicability (low/ high/ unclear) to your review question defined above.

*Complete all domains separately for each evaluation of a distinct model. Shaded boxes indicate where signalling questions do not apply and should not be answered.*

| DOMAIN 1: Participants | | | |
|---|---|---|---|
| **A. Risk of Bias** | | | |
| *Describe the sources of data and criteria for participant selection:*<br>Source of data is hospitals in Spain, the US and Southern American countries. All patients with diagnosed COVID-19 during a given a period of time were included. Variables were selected based on missingness and common variables between different cohorts. | | | |
| | | Dev | Val |
| 1.1 Were appropriate data sources used, e.g. cohort, RCT or nested case-control study data? | | Yes | Yes |
| 1.2 Were all inclusions and exclusions of participants appropriate? | | Yes | Yes |
| **Risk of bias introduced by selection of participants** | **RISK:**<br>*(low/ high/ unclear)* | **Low** | **Low** |
| *Rationale of bias rating:*<br>All participants from the hospitals were included. | | | |
| **B. Applicability** | | | |
| *Describe included participants, setting and dates:*<br>Figure 1 and pages 5, 11 and 14 of main text. | | | |
| **Concern that the included participants and setting do not match the review question** | **CONCERN:**<br>*(low/ high/ unclear)* | **Low** | **Low** |
| *Rationale of applicability rating:*<br>All participant charcateristics were indicated along the manuscript. | | | |

| DOMAIN 2: Predictors | | | |
|---|---|---|---|
| **A. Risk of Bias** | | | |
| ***List and describe predictors included in the final model, e.g. definition and timing of assessment:***<br>Age (years), Platelet Count (x 10^6/L), Eosinophils (x 10^6/L), Neutrophils (x 10^6/L), Monocytes (x 10^6/L), C-Reactive Protein (mg/L), Creatinine (mg/dL), Lactate Dehydrogenase (U/L), Serum Sodium (mmol/L), Serum Potassium (mmol/L), Glucose (mg/dL) and Dimer (ng/mL). From the 5th February 2020 to the 7th June 2021. | | | |
| | | Dev | Val |
| 2.1 Were predictors defined and assessed in a similar way for all participants? | | Yes | Yes |
| 2.2 Were predictor assessments made without knowledge of outcome data? | | Yes | Yes |
| 2.3 Are all predictors available at the time the model is intended to be used? | | Yes | Yes |
| **Risk of bias introduced by predictors or their assessment** | **RISK:**<br>*(low/ high/ unclear)* | **Low** | **Low** |
| *Rationale of bias rating:*<br>Predictors are normal blood test values and age. | | | |
| **B. Applicability** | | | |
| Concern that the definition, assessment or timing of predictors in the model do not match the review question | **CONCERN:**<br>*(low/ high/ unclear)* | **Low** | **Low** |
| ***Rationale of applicability rating:***<br>We have considered different timings from the first day at hospital until 9th day at hospital. | | | |

| DOMAIN 3: Outcome | | | |
|---|---|---|---|
| **A. Risk of Bias** | | | |
| ***Describe the outcome, how it was defined and determined, and the time interval between predictor assessment and outcome determination:***<br>Outcome was the status of subject when leaving the hospital: either death or alive. | | | |
| | | Dev | Val |
| 3.1 Was the outcome determined appropriately? | | Yes | Yes |
| 3.2 Was a pre-specified or standard outcome definition used? | | Yes | Yes |
| 3.3 Were predictors excluded from the outcome definition? | | Yes | Yes |
| 3.4 Was the outcome defined and determined in a similar way for all participants? | | Yes | Yes |
| 3.5 Was the outcome determined without knowledge of predictor information? | | Yes | Yes |
| 3.6 Was the time interval between predictor assessment and outcome determination appropriate? | | Yes | Yes |
| **Risk of bias introduced by the outcome or its determination** | **RISK:**<br>*(low/ high/ unclear)* | **Low** | **Low** |
| ***Rationale of bias rating:***<br>Risk of bias in determining the defined outcome is very low. | | | |
| **B. Applicability** | | | |
| ***At what time point was the outcome determined:***<br>At the time (number of days) when subject left the hospital.<br><br>***If a composite outcome was used, describe the relative frequency/distribution of each contributing outcome:***<br>n/a | | | |
| **Concern that the outcome, its definition, timing or determination do not match the review question** | **CONCERN:**<br>*(low/ high/ unclear)* | **Low** | **Low** |
| ***Rationale of applicability rating:***<br>We considered over survival and thus concern is low. | | | |

| DOMAIN 4: Analysis |
|---|
| **Risk of Bias** |

***Describe numbers of participants, number of candidate predictors, outcome events and events per candidate predictor:***
Figure 1 and Supplementary Tables 1-4.

***Describe how the model was developed (for example in regards to modelling technique (e.g. survival or logistic modelling), predictor selection, and risk group definition):***
The model was developed using iterative variable selection and linear LASSO model. Risk groups were selected in three different ways: Youden's J statistics, threshold from training cohort to equal 95% sensitivity and 95 specificity.

***Describe whether and how the model was validated, either internally (e.g. bootstrapping, cross validation, random split sample) or externally (e.g. temporal validation, geographical validation, different setting, different type of participants):***
We used both internal validation (10-fold cross-validation in model building) and different external validations (temporal and geographical external validation, different ealthcare systems).

***Describe the performance measures of the model, e.g. (re)calibration, discrimination, (re)classification, net benefit, and whether they were adjusted for optimism:***
We used AUROC, accuracy, sensitivity, specificity, horizon analysis at 9 days before clinical resolution, and survival analysis. We checked calibration for all cohorts.

***Describe any participants who were excluded from the analysis:***
Participants with obvious outlier values (e.g. hospitalisation date after date of outcome) and with no PCR information, were excluded.

***Describe missing data on predictors and outcomes as well as methods used for missing data:***
Missing values in predictors were imputed using training cohort mean values. There were no missing values in outcomes.

| | | | Dev | Val |
|---|---|---|---|---|
| 4.1 | Were there a reasonable number of participants with the outcome? | | Yes | Yes |
| 4.2 | Were continuous and categorical predictors handled appropriately? | | Yes | Yes |
| 4.3 | Were all enrolled participants included in the analysis? | | Yes | Yes |
| 4.4 | Were participants with missing data handled appropriately? | | Yes | Yes |
| 4.5 | Was selection of predictors based on univariable analysis avoided? | | Yes | |
| 4.6 | Were complexities in the data (e.g., censoring, competing risks, sampling of controls) accounted for appropriately? | | Yes | Yes |
| 4.7 | Were relevant model performance measures evaluated appropriately? | | Yes | Yes |
| 4.8 | Were model overfitting and optimism in model performance accounted for? | | Yes | |
| 4.9 | Do predictors and their assigned weights in the final model correspond to the results from multivariable analysis? | | Yes | |
| **Risk of bias introduced by the analysis** | | **RISK:** *(low/ high/ unclear)* | **Low** | **Low** |

*Rationale of bias rating:*
All aforementioned steps in the modelling were considered.

**Step 4: Overall assessment**

Use the following tables to reach overall judgements about risk of bias and concerns regarding applicability of the prediction model evaluation (development and/or validation) across all assessed domains.
*Complete for each evaluation of a distinct model.*

| **Reaching an overall judgement about risk of bias of the prediction model evaluation** | |
|---|---|
| **Low risk of bias** | If all domains were rated low risk of bias.<br>If a prediction model was developed without any external validation, and it was rated as low risk of bias for all domains, consider downgrading to **high risk of bias**. Such a model can only be considered as low risk of bias, if the development was based on a very large data set and included some form of internal validation. |
| **High risk of bias** | If at least one domain is judged to be at **high risk of bias**. |
| **Unclear risk of bias** | If an unclear risk of bias was noted in at least one domain and it was low risk for all other domains. |

| **Reaching an overall judgement about applicability of the prediction model evaluation** | |
|---|---|
| **Low concerns regarding applicability** | If low concerns regarding applicability for all domains, the prediction model evaluation is judged to have **low concerns regarding applicability**. |
| **High concerns regarding applicability** | If high concerns regarding applicability for at least one domain, the prediction model evaluation is judged to have **high concerns regarding applicability**. |
| **Unclear concerns regarding applicability** | If unclear concerns (but no "high concern") regarding applicability for at least one domain, the prediction model evaluation is judged to have **unclear concerns regarding applicability** overall. |

| **Overall judgement about risk of bias and applicability of the prediction model evaluation** | | |
|---|---|---|
| **Overall judgement of risk of bias** | **RISK:**<br>*(low/ high/ unclear)* | **Low** |
| *Summary of sources of potential bias:*<br>All domains were rated low risk of bias. | | |
| **Overall judgement of applicability** | **CONCERN:**<br>*(low/ high/ unclear)* | **Low** |
| *Summary of applicability concerns:*<br>All domains were rated low risk of bias. Furthermore, the predictor was externally validated in 4 different countries. | | |
